# Workload Indicators of Staffing Need (WISN) Method for Midwives Planning and Estimation at Asrade Zewude Memorial Primary Hospital, North west Ethiopia

**DOI:** 10.1101/2022.03.28.22273061

**Authors:** Gizew Dessie Asres

## Abstract

**Background:** Work force is a crucial component of health services delivery system. Ethiopia faces health workforce challenges with regard to evidence based health work force planning. First the health worker to population ratio was used and later standard staffing schedule for each health facility level was used. Both of these methods did not address the issue of evidence based workload variation between the same levels of facilities at different locations within a country. A workload indicator of staffing need method (WISN) addresses these variations. Therefore this study was assumed for recommendation on use of WISN for health facilities based on WISN results of midwives at Asrade Zewude memorial Hospital.

**Methods:** Cross sectional study using WISN model was used to determine the gaps or excess and workload pressure in midwives at Asrade Zewude Memorial primary hospital, North West Ethiopia.

**Results:** The finding showed that there was five working days with 1030 hrs actual working time per year for midwives. This working time was spent on health service activities (58.4%), additional activities (36.6%) and support activities (5%). WISN calculation showed that a shortage of five midwives with WISN ratio of 0.8 at Asrade Zewude Memorial primary hospital North West Ethiopia.

**Conclusion:** Midwives at the study area were doing their routine work with 20% under staffed by covering the additional five midwives’ position. With this working condition, it may be hard to achieve universal health coverage goals of the facility. Therefore the hospital should institutionalize WISN method to objectively employ midwifery professionals.

## Introduction

Globally countries at all levels of socioeconomic development face, to varying degrees, difficulties in the education, deployment, retention, and performance of their health workforce. Health priorities of the post-2015 agenda for sustainable development will remain inspirational unless accompanied by strategies involving transformational efforts on competent health workforce planning and deployment [1-3].

Countries have been planning for human resources for health for a long time. In parallel, demand has grown for tools to facilitate that planning, including tools that can assist with applying objective and scientific methodologies to estimate health workforce requirements [4-6]. In order to improve health care delivery, health services’ managers are faced with increasing challenges in providing adequate and quality health workforce to meet the demand of the ever growing population. Human resource for health (HRH) is one of the six building blocks of the WHO framework for health systems. Availability of trained health workers is one of the most critical limiting factors for the delivery of a minimum health service package [7, 8].

There is an increasing need for health organizations to identify the most appropriate staffing levels and skill mix to ensure efficient and effective use of the limited resources. Often this is not the case as the required cadres are frequently missing in specific geographical areas or health facilities where they are needed most; or in surplus where need is low resulting into inefficiencies [9]. Poor planning and management of human resource for health among other factors, contribute to these shortages. The required number of health workers and their skill mix in a health facility will depend on the workload and the range of services in the facility which in most cases is related to the minimum health care package of the facility [4, 10-11].

Application of Workload Indicators of Staffing Need (WISN) method in planning and projection of human resources for health helps to rectify many of the observed deficiencies in access to human resource for health. This implies each health facility has its own staffing requirement depending on its workload. Following its development by WHO, the WISN has been used to determine staffing requirements in Papua New Guinea, the United Republic of Tanzania, Kenya, Sri Lanka and also in six other countries: Bahrain, Egypt, Hong Kong, Oman, Sudan and Turkey among others [4, 7, 12-21].

Ethiopia is facing challenges in ensuring human resources for health: availability, accessibility, acceptability, quality and effective coverage due to mainly poor human resources for health management. The health worker to population ratio method and standard staffing schedule for each health facility level are no longer effective for health worker planing. Therefore the WISN method of staff requirement to health facilities should be advocated as an option by applying it at Asrade Zewude Memorial primary hospital.

## Methods

Facility based cross sectional design was done using WISN model to compare available midwives with required midwives. So that the gaps or excess and workload pressure of midwives were calculated at Asrade Zewude Memorial primary hospital, North West Ethiopia. Midwives were selected as priority cadre based on current staffing problems. Available working time was determined using national calendar, personnel files and HR regulatory documents. Workload components of midwives were determined by establishing expert working group comprised of midwives hospital performance monitoring and evaluation unit staff and human resources management staff. Two hours detail training was given for expert working group using WISN user manual. The data that has been used by expert working group were; detail midwives job descriptions obtained from human resources management, annual service statistics obtained from hospital performance monitoring and evaluation unit and midwifery service activities and support activities were listed and explained by head of hospital midwifery service unit. The workload data were cross checked from July 1, 2019–June 30, 2020 G.C for midwifery department using health service activity data of district health information system (DHS2) data base.

The WISN tool takes into account certain information in calculating staffing requirements for health service delivery points. The required information includes the common activities performed by midwives on a day-to-day basis at Asrade Zewude Memorial Primary Hospital (i.e., workload components), the time it takes a midwife to conduct core activities and associated activities (i.e., activity standards), working time available in 1 year for a midwife to execute their work and the annual service delivery statistics in the selected health service delivery point [11, 12].

### Determining priority cadres

The WISN method can be applied to all health worker cadres and all types of health facilities. It is unlikely, however, that we have not sufficient resources to do it all at one time. We thus decided which staff categories working in the hospital would be our WISN target (see Table 1). It is generally better to start with the cadres with less complex activity standards to list. We can then expand this to other cadres in subsequent WISN applications, after our team gain experience with the method.

**Table 1.** Determining priority health worker categories for WISN study at Asrade Zewude Primary hospital, 2020

| Work unit | Staff category | Staffing problems (current and likely in the future) | Priority point scale 0 to 10 |
| --- | --- | --- | --- |
| Emergency | Doctors | We have adequate number of day time Doctors | 0 |
|  | Integrated Emergency Surgical Officer(IESO) | Small, but not serious, shortage of IESOs in the hospital in the Emergency during working hours | 5 |
|  | Nurses | Small, but not serious, shortage of Nurses in the hospital in the Emergency during working hours | 6 |
|  | Anesthetist | Small, but not serious, shortage of Anesthetists in the hospital in the Emergency during working hours | 4 |
| IPD | Doctors | We have adequate number of day time Doctors | 2 |
|  | Integrated Emergency Surgical Officer | Small, but not serious, shortage of IESOs in the hospital in the inpatient ward during working hours | 5 |
|  | Nurses | Small and serious, shortage of Nurses in the hospital in the inpatient wards | 7 |
|  | Mid wives | Small and serious, shortage of midwives in the hospital in the inpatient wards | 9 |
|  | Laboratory | Small, but not serious, shortage of laboratory professionals in the hospital in the inpatient ward during working hours | 7 |
|  | Pharmacy | Small, but not serious, shortage of pharmacy professionals in the hospital in the inpatient ward during working hours | 7 |
|  | Anesthetist | Small, but not serious, shortage of Anesthetists in the hospital in the inpatient ward during working hours | 4 |
| OR | Integrated Emergency Surgical Officer | Small, but not serious, shortage of IESOs in the hospital in the OR during working hours | 5 |
|  | Doctors | We have adequate number of day time Doctors | 3 |
|  | Nurses | Small, but not serious, shortage of Nurses in the hospital in the OR during working hours | 5 |
|  | Midwives | Small and serious, shortage of midwives in the hospital in the OR | 9 |
|  | Anesthetist | Small, but not serious, shortage of Anesthetists in the hospital in the inpatient ward during working hours | 4 |
| OPD | Doctors | We have adequate number of Doctors | 5 |
|  | Integrated Emergency Surgical Officer | Small, but not serious, shortage of IESOs in the hospital in the OPD | 5 |
|  | Nurses | Small, but not serious, shortage of Nurses in the hospital in the OPD during | 7 |
|  | Mid wives | Small and serious, shortage of midwives in the hospital in the OPD | 10 |
|  | Laboratory | Small, but not serious, shortage of laboratory professionals in the hospital in the OPD | 7 |
|  | Pharmacy | Small, but not serious, shortage of pharmacy professionals in the hospital in the OPD | 7 |
|  | Radiographers | Small, but not serious, shortage of radiographer in the hospital in the OPD | 4 |
| NICU | Doctors | We have adequate number of day time Doctors |  |
|  | Integrated Emergency Surgical Officer | Small, but not serious, shortage of IESOs in the hospital in the NICU | 5 |
|  | Nurses | Small but not serious, shortage of nurses in the hospital | 8 |
|  | Mid wives | Small but not serious, shortage of midwives in the hospital | 5 |
| Labor and delivery | Midwives | Small but not serious, shortage of midwives in the hospital | 9 |
|  | Integrated Emergency Surgical Officer | Small, but not serious, shortage of IESOs in the hospital | 5 |
|  | Doctors | We have adequate number of Doctors | 1 |
| **Highest Priority score cadres: 1. Midwives first highest priority. 2. Nurses second highest priority. 3. Laboratory and pharmacy third highest priority.** |  |  |  |

We set our priorities in a systematic way. First, we have listed all work units (as appropriate) and the main staff categories working here. Second, we determined which are our most difficult staffing problems regarding these staff cadres. We wrote them down. We consider our current staffing problems, as well as those that we anticipate having in the future. Third, we decided which staff category (or categories) should have highest priority. Here are some questions to consider in making our selection:

- Which staff category is in shortest supply in relation to the need for staff?
- Which of these staffing problems have affected the quality of care most?
- Which of them are likely to affect the quality of care soon?
- Are any of the staff cadres particularly important for planned future health programs?

## Result

Based on this study the current staff assigned using standard staffing schedule (fixed number of health worker for the same level of health facility, locally called form 15) and the required staff based on current service standard showed discrepancy. Number of midwives during data collection was 20 but the required number of midwives based on WISN calculation was 25, only 80% of the staff needed was available.

The working days per week were five days and the working hours per day was 7.8. The annual and other leaf was 25 days /year and total annual working time was 1030 hours (see **Table 2**).

**Table 2.** Available Working Time (AWT) for Midwives at Asrade Zewude Memorial primary hospital, 2020 G.C.

| Working Days Per Week (A) | Working Hours Per Day (B) | Annual and other Leaves days(C) | Public Holidays (D) | Training Days Per Year (E) | AWT In Weeks Per Year (F=A*B) | Total working days per year(G) | Awt In Days Per Year (H=G-(C+D+E)) | Awt In Hours Per Year (I=H*B) |
| --- | --- | --- | --- | --- | --- | --- | --- | --- |
| 5 | 7.8(39/5) | 25 days | 6 | 82 | 49 | 245 | 132 | 1030 |

Source: Asrade Zewude Memorial Hospital Human resource department

Activity standards for midwives working at Asrade Zewude Memorial Hospital were fifteen health service activities, four support activities and five additional activities (see Table 3).

**Table 3.** Activity standards for midwives working at Asrade Zewude Memorial Hospital, 2020 G.C.

| Workload group | Workload component |
| --- | --- |
| Health service activities of all midwives | 1. Cervical cancer screening |
|  | 2. Short term family planning service |
|  | 3. Long term family planning service |
|  | 4. First Ante natal care |
|  | 5. fourth Ante natal care |
|  | 6. labour with partograph without episotomy |
|  | 7. Labour with phartograph with Episotomy |
|  | 8. Preparing laboring mothers to Cesarian section/ OR |
|  | 9. Post Natal Care |
|  | 10. Recovery |
|  | 11. Safe Abortion |
|  | 12. Post abortion care |
|  | 13. Immunization |
|  | 14. High risk mothers care from admission to follow up per day |
|  | 15. Birth notification |
| Support activities of all midwives | 1. Documentation to registration and Tally 10min per client |
|  | 2. Meetings 1hrs/2 weeks |
|  | 3. Health center mentorship 8hrs 2 midwives/16 health center/2month |
|  | 4. Accompanying reoffered out laboring mothers 8hrs/client |
| Additional activities of certain midwives | 1. Participating Management meeting 3hrs/week |
|  | 2. Schedule posting or task assignment of midwives 30hrs/month |
|  | 3. Reporting performances of the unit 8hrs/month |
|  | 4. Evaluating performances of each mid wife/PA 5days/6month |
|  | 5. Reporting monthly attendances for HRM unit 10min/month |

Source: Asrade Zewude Memorial Hospital midwifery department

Based on expert working group observation, attending labor with partograph and doing episiotomy took 12 hrs. Attending labor with partograph without episiotomy took 8 hrs. Health service activities that took shorter time were family planning, preparing mothers for cesarean section, immunization at birth, postnatal care and birth notification (see Table 4)

**Table 4.**
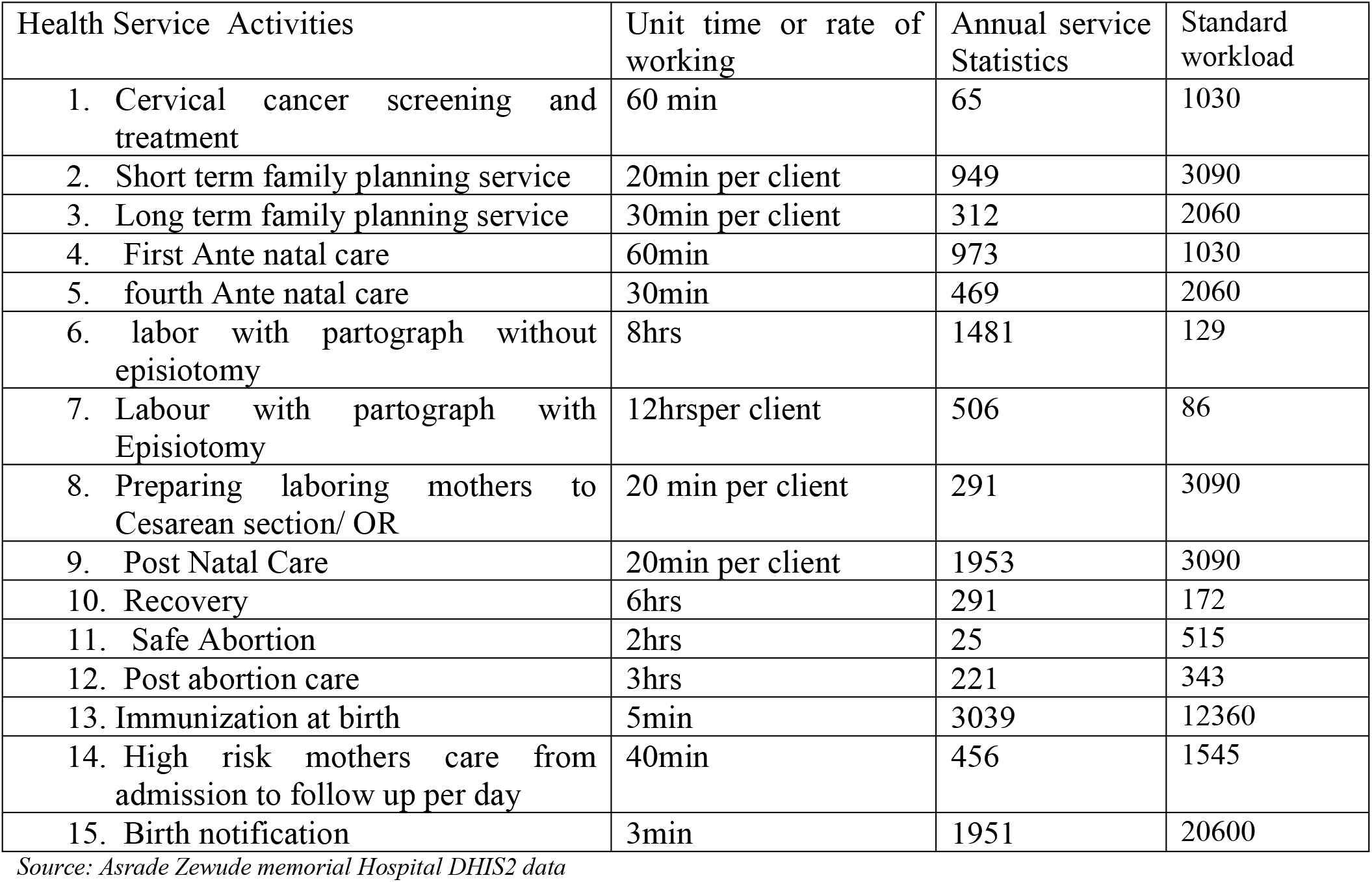
Service Standards and Annual Service Statistics of Midwifery Related Activities at Asrade Zewude Memorial Primary Hospital, 2020 G.C.

| Health Service Activities | Unit time or rate of working | Annual service Statistics | Standard workload |
| --- | --- | --- | --- |
| 1. Cervical cancer screening and treatment | 60 min | 65 | 1030 |
| 2. Short term family planning service | 20min per client | 949 | 3090 |
| 3. Long term family planning service | 30min per client | 312 | 2060 |
| 4. First Ante natal care | 60min | 973 | 1030 |
| 5. fourth Ante natal care | 30min | 469 | 2060 |
| 6. labor with partograph without episiotomy | 8hrs | 1481 | 129 |
| 7. Labour with partograph with Episiotomy | 12hrsper client | 506 | 86 |
| 8. Preparing laboring mothers to Cesarean section/ OR | 20 min per client | 291 | 3090 |
| 9. Post Natal Care | 20min per client | 1953 | 3090 |
| 10. Recovery | 6hrs | 291 | 172 |
| 11. Safe Abortion | 2hrs | 25 | 515 |
| 12. Post abortion care | 3hrs | 221 | 343 |
| 13. Immunization at birth | 5min | 3039 | 12360 |
| 14. High risk mothers care from admission to follow up per day | 40min | 456 | 1545 |
| 15. Birth notification | 3min | 1951 | 20600 |

Source: Asrade Zewude memorial Hospital DHIS2 data

The support activities account 5% of the total working hours of which 3% spend on meeting (see Table 5).

**Table 5.** Support Activities of Midwifery staff at Asrade Zewude Memorial Primary Hospital, 2020 G.C.

| Workload Components | CAS (actual working time) | CAS % (percentage working time) |
| --- | --- | --- |
| 1. Documentation to registration and Tally | 10min/day | 2% = $(10/60)/7.8 \times 100$ |
| 2. Meetings | 2hrs/2weeks | 3% = $2\text{hrs}/(7.8\text{hrs} \times 5\text{days} \times 2\text{wks}) \times 100$ |
| Total CAS % |  | 5% |

Source: Asrade Zewude Memorial Hospital Human resource and midwifery department

Midwives at Asrade Zewude Memorial Primary Hospital spent a total of 377 hrs. in a year for additional activities. Additional activities which took longer time were evaluating or performance appraisal of the unit, health center mentorship and preparing performance report. While reporting monthly attendances and schedule posting took shorter time (see **Table 6)**

**Table 6.**
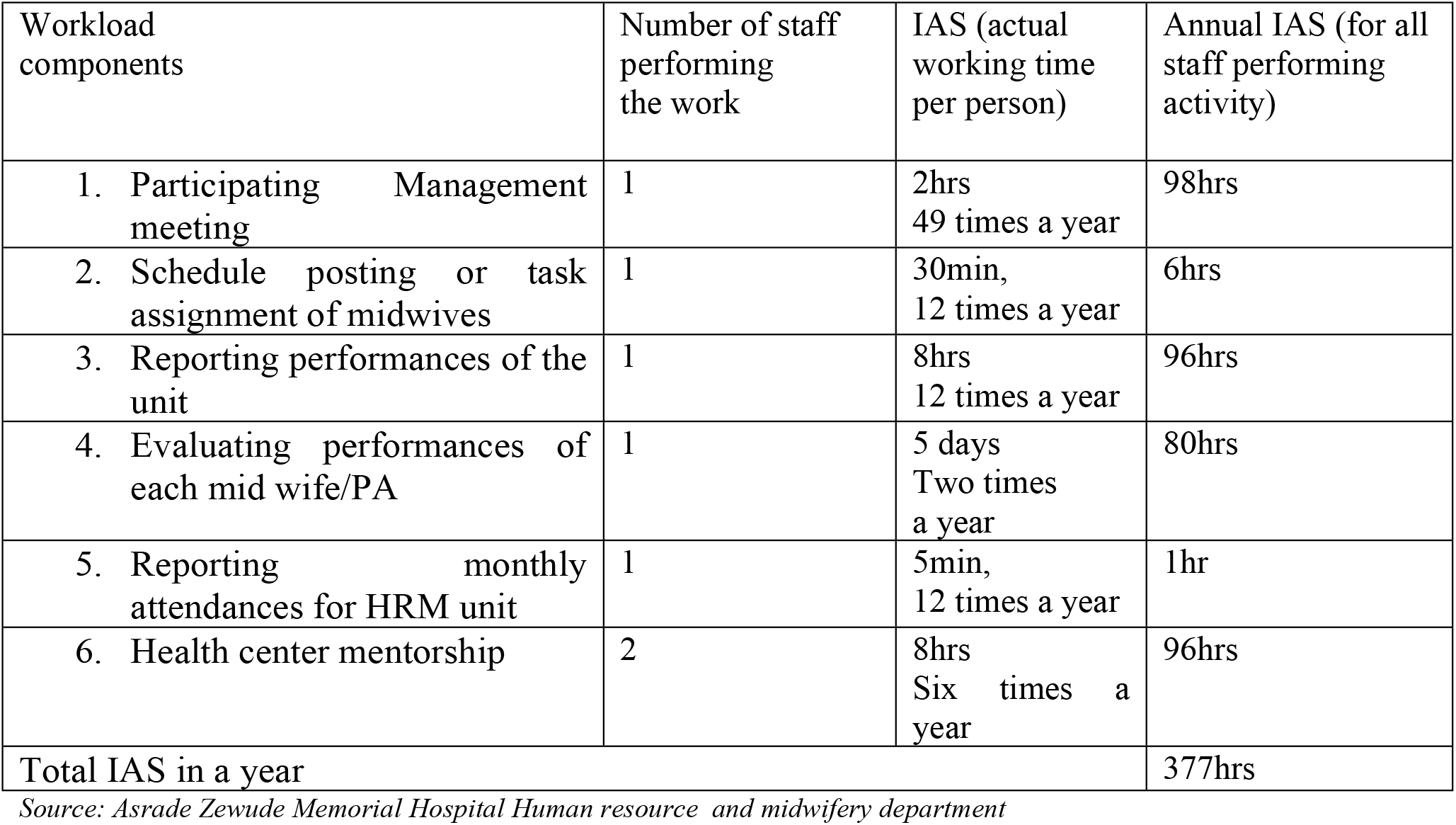
Additional Activities of Certain Midwives at Asrade Zewude Memorial Primary Hospital, 2020 G.C

| Workload components | Number of staff performing the work | IAS (actual working time per person) | Annual IAS (for all staff performing activity) |
| --- | --- | --- | --- |
| 1. Participating Management meeting | 1 | 2hrs<br>49 times a year | 98hrs |
| 2. Schedule posting or task assignment of midwives | 1 | 30min,<br>12 times a year | 6hrs |
| 3. Reporting performances of the unit | 1 | 8hrs<br>12 times a year | 96hrs |
| 4. Evaluating performances of each mid wife/PA | 1 | 5 days<br>Two times a year | 80hrs |
| 5. Reporting monthly attendances for HRM unit | 1 | 5min,<br>12 times a year | 1hr |
| 6. Health center mentorship | 2 | 8hrs<br>Six times a year | 96hrs |
| Total IAS in a year |  |  | 377hrs |

*Source: Asrade Zewude Memorial Hospital Human resource and midwifery department*

Calculation based on WISN showed that about twenty five midwives were needed for health service activities support activities and additional activities (see **Table 7**).

**Table 7.** Determining staff requirements based on WISN for Midwives at Asrade Zewude Memorial Primary Hospital, 2020 G.C

| Health service activities of all cadre members | Workload Component | Annual Workload | Standard Workload | Required number of staff members |
| --- | --- | --- | --- | --- |
|  | 1. Cervical cancer screening | 65 | 1030 | 0.63 |
|  | 2. Short term family planning service | 949 | 3090 | 0.31 |
|  | 3. Long term family planning service | 312 | 2060 | 0.15 |
|  | 4. First Ante natal care | 973 | 1030 | 0.94 |
|  | 5. fourth Ante natal care | 469 | 2060 | 0.23 |
|  | 6. labor with pantograph without episiotomy | 1481 | 129 | 11.48 |
|  | 7. Labour with partograph with Episiotomy | 506 | 86 | 5.88 |
|  | 8. Preparing laboring mothers to Cesarean section/ OR | 291 | 3090 | 0.09 |
|  | 9. Post Natal Care | 1953 | 3090 | 0.63 |
|  | 10. Recovery | 291 | 172 | 1.69 |
|  | 11. Safe Abortion | 25 | 515 | 0.05 |
|  | 12. Post abortion care | 221 | 343 | 0.64 |
|  | 13. Immunization | 3039 | 12360 | 0.25 |
|  | 14. High risk mothers care from admission to follow up per day | 456 | 1545 | 0.29 |
|  | 15. Birth notification | 1951 | 20600 | 0.09 |
| **A. Total required staff for health service activities** |  |  |  | **23.35** |
| Support activities of all cadre members | Workload Component | CAS (Actual working time) | CAS (Percentage working time) |  |
|  | Documentation to registration and Tally | 10min/day | 2%=(10/60)/7.8*100 |  |
|  | Meetings | 2hrs/2weeks | 3%=2hrs/(7.8hrs*5days*2wks))*100 |  |
| Total CAS % |  |  | 5% |  |
| **B. Category allowance factor:** $\{1 / [1 - (\text{total CAS percentage} / 100)]\} = 1.05$ | | | | |
| Additional activities of certain cadre members | Workload Component | Number of staff members performing the work | IAS (Actual working time per person) | Annual IAS (for all staff performing activity) |
|  | Participating Management meeting | 1 | 2hrs<br>49 times a year | 98hrs |
|  | Schedule posting or task assignment of midwives | 1 | 30min,<br>12 times a year | 6hrs |
|  | Reporting performances of the unit | 1 | 8hrs<br>12 times a year | 96hrs |
|  | Evaluating performances of each mid wife/PA | 1 | 5 days<br>Two times a year | 80hrs |
|  | Reporting monthly attendances for HRM unit | 1 | 5min,<br>12 times a year | 1hr |
|  | Health center mentorship | 2 | 8hrs<br>Six times a<br>year | 96hrs |
| Total IAS in a year |  |  |  | 377hrs |
| **C. Individual allowance factor (Annual total IAS / AWT)** |  |  |  | 0.37 |
| **Total required number of staff based on WISN: (A x B + C)** |  |  |  | 24.89 (□25) |

*Source: Asrade Zewude Memorial Hospital Human resource, midwifery department and survey analysis*

## Discussion

This study demonstrated the implementation process of the WISN methodology using midwifery staff data at Asrade Zewude Memorial Hospital. According to this study, the midwives at Asrade Zewude Memorial Hospital had fifteen workload components under health service activity, six workload components under additional activity and two workload components as support activity.

The finding showed that there was five working days with 1030 hrs actual working time per year for midwives at Asrade Zewude Memorial Hospital. This working time was spent on health service activities (58.4%), additional activities (36.6%) and support activities (5%).

According to the study, the traditional way of staffing method, actual number of midwives and WISN method has varied result. Based on Form 15 (fixed number of health worker for the same level of health facility) the number of midwives expected was 8 and actual number of midwives was 20 but when calculated based on available data 25 midwives were needed based on WISN method.

The workload pressure in the study area was found to be WISN ratio of 0.8. This was higher than a study done in Uganda (0.58), India (0.6), Greek (1.33-1.83), Bangladesh(0.69) and Indonesia (0.64) [22-26].

## Conclusion

WISN method is useful in evidence based estimating of staff requirements based on available evidences. It enables health facilities to staff with adequate health workers based on objective workload evidence. WISN method of workforce planning had challenges related to proper documentation and data quality. Therefore institutionalizing this tool enables health facilities to focus and strengthen data quality, another dimension of the health system.

Based on the WISN output of Asrade Zewude memorial Hospital the following recommendation was forwarded to stakeholders.

➢ Decision makers at regional health bureau and federal ministry of health should better institutionalize WISN method to have the right number of midwives at the right time.
➢ Human resource department should advocate the use of WISN in the health facility.
➢ The planning and health information department should enhance WISN based planning as it encourages performance and data quality.

## Data Availability

All necessary data are provided in the manuscript.

## Declarations

### Ethics approval and consent to participate

NA

### Consent for publication

Consent for publication was obtained from Asrade zewude Hospital CEO

### Availability of data and materials

Necessary materials may be available upon request.

### Authors’ contributions

G.D.A did from conceptualization to writing up the whole research document and analysis

## Acknowledgements

I would like to appreciate the contributions made by the midwifery department staff and Human Resource staff at Asrade Zewude Hospital.

